# Development and Validation of a Simple Risk Score for Diagnosing COVID-19 in the Emergency Room

**DOI:** 10.1101/2020.08.11.20173112

**Authors:** Joowhan Sung, Naveed Choudry, Rima Bachour

**Author notes:** Address for Correspondence: Joowhan Sung, MD, Georgetown Hospitalist Office, MedStar Southern Maryland Hospital, 7503 Surratts Rd, Clinton, MD 20735, USA.

## Abstract

As the COVID-19 pandemic continues to escalate and place pressure on hospital system resources, a proper screening and risk stratification score is essential. We aimed to develop a risk score to identify patients with increased risk of COVID-19, allowing proper identification and allocation of limited resources. A retrospective study was conducted of 338 patients who were admitted to the hospital from the emergency room and tested for COVID-19 at an acute care hospital in the Metropolitan Washington D.C. area. The dataset was split into development and validation sets with a ratio of 6:4. Demographics, presenting symptoms, sick contact, triage vital signs, initial laboratory and chest X-ray results were analyzed to develop a prediction model for COVID-19 diagnosis. Multivariable logistic regression was performed in a stepwise fashion to develop a prediction model, and a scoring system was created based on the coefficients of the final model. Among 338 patients admitted to the hospital from the emergency room, 136 (40.2%) patients tested positive for COVID-19 and 202 (59.8%) patients tested negative. Nursing facility residence (2 points), sick contact (2 points), constitutional symptom (1 point), respiratory symptom (1 point), gastrointestinal symptom (1 point), obesity (1 point), hypoxia at triage (1 point), and leukocytosis (−1 point) were included in the prediction score. A risk score for COVID-19 diagnosis achieved AUROC of 0.87 (95% CI 0.83-0.92) in the development dataset and 0.83 (95% CI 0.76-0.90) in the validation dataset. A risk prediction score for COVID-19 can be used as a supplemental tool to assist clinical decision to triage, test, and quarantine patients admitted to the hospital from the emergency room.

## Introduction

In December of 2019, an outbreak of a novel coronavirus disease was reported in the Hubei Province of China. Caused by the emerging virus Severe Acute Respiratory Syndrome Coronavirus 2 (SARS-CoV-2), the disease has quickly spread across the world. On January 30th, 2020 the World Health Organization (WHO) declared the outbreak a Public Health Emergency of International Concern, and on March 11th, 2020, declared it a pandemic [1]. As of Aug 6th, 2020, there are over 19.0 million confirmed cases worldwide, with 4.8 million being in the United States of America (USA) [2].

The pandemic has caused significant adverse impacts throughout the USA and the world. Since the early days of the COVID-19 pandemic in the USA, hospital systems have found themselves overwhelmed and with limitations in capacity to triage, diagnose and treat patients afflicted by COVID-19. As the pandemic continues unabated and as it spreads through the United States, it is necessary to improve hospital screening and stratification of at-risk populations to enable timely and appropriate quarantine, treatment, and use of limited resources.

At present, no validated risk score or stratification system is readily available to aid the clinical decisionmaking process of hospital-based staff in determining when testing for COVID-19 is appropriate [3]. Availability of testing for COVID-19 continues to be an ongoing limitation throughout the United States. A system of clinical risk stratification can help to identify patients that present a higher risk and warrant COVID-19 testing in a resource limited setting.

In this retrospective study, we reviewed the records of patients presenting to an emergency department in an acute care hospital in the Metropolitan Washington D.C. area who were tested for SARS-CoV-2 and admitted. We reviewed the clinical characteristics, radiographic findings, and laboratory findings between those who tested positive and negative, then developed a simple bedside scoring system based on a risk prediction score.

## Methods

### Cohort design and subjects

A retrospective review was performed for patients tested for COVID-19 and admitted to MedStar Southern Maryland Hospital, a 262-bed acute care hospital located in a suburb of Washington D.C. between April 1, 2020, and April 30, 2020. During this time, the hospital and surrounding region experienced a surge of COVID-19 admissions, but universal testing for COVID-19 was not performed for hospitalized patients. Patients were included in the study if they presented to the emergency room and were admitted to the hospital with laboratory-confirmed COVID-19 (cases) or tested negative for COVID-19 within 24 hours of hospital admission (controls). All COVID-19 diagnosis was made by nasopharyngeal swab and RT-PCR for SARS-CoV-2. Patients who were admitted from the emergency room directly to the intensive care unit (ICU) were excluded from the study. These patients were excluded as a majority of them were not in a condition to describe their symptoms at the time of the presentation, and the severity of their symptoms often necessitated COVID-19 testing during empiric work up.

### Data collection

For all eligible patients, records from initial hospital encounter were reviewed. Demographics (age, sex, race, and smoking status), past medical history (diabetes, hypertension, chronic obstructive pulmonary disease [COPD], asthma, coronary artery disease [CAD] congestive heart failure [CHF], atrial fibrillation, chronic kidney disease [CKD], and end-stage renal disease [ESRD]), sick contact with suspected COVID-19 case, presenting symptoms (fever, chills, myalgia, cough, shortness of breath, nausea, vomiting or diarrhea), triage vital signs (temperature, heart rate, systolic and diastolic blood pressure, respiratory rate, and oxygen saturation), initial basic laboratory test results (complete blood count, and creatinine) and chest X-ray results were collected.

### Development of prediction model

The dataset was randomly split into a development cohort and a validation cohort with a 6:4 ratio using function of statistical software. Baseline characteristics were compared between cases and controls within each cohort. Categorical variables were compared using the chi-squared test and continuous variables were compared using the student t-test. Univariable logistic regression was performed in a development cohort to identify potential predictors of COVID-19 status. Variables associated with COVID-19 status (p<0.1) became candidates for a multivariable model. A multivariable logistic regression model was built in a stepwise fashion. Variables identified in univariable analysis entered the model one by one and retained in the model if the addition of the variable improved the fit of the model. Variables with p-value higher than 0.1 were removed from the model. A risk scoring system was created based on the coefficients from the final logistic regression model. The risk score was validated in the testing cohort. The area under the receiver operating curve was calculated. The analysis was performed using STATA version 15.1 (STATA Corp., Texas, USA).

### Ethical consideration

This study was approved by the Institutional Review Board (IRB) of the MedStar Health Research Institute with a waiver of individual consents. (IRB ID: MOD00004296)

## Results

### Study population

A total of 656 patients were admitted to the hospital during the study period. Of them, 79 patients who were admitted to the intensive care unit directly from the emergency room were excluded from the study. Among 577 patients admitted to the medical floor, 338 patients received testing for COVID-19 and were included in this study. Of those included, 136 (40.2%) patients tested positive for SARS-CoV-2, and 202 (59.8%) patients tested negative. Demographic characteristics of patients were described in Table 1. In the entire cohort, the median age was 65 years old (interquartile range [IQR] 54-76 years old), 53% were males, 82.8% were African Americans, 10.1% were Hispanics and 5.6% were Caucasians, 30.5% were current or former smokers, 14.2% were from skilled nursing facility, 42.9% were obese, 64.5% had hypertension, 38.5% had diabetes, 21.0% had CKD, 16.0% had CHF, and 12.7% had COPD.

**Table 1.** Baseline characteristics of the patients admitted to the hospital in a development and a validation cohort.

|  | Development cohort<br>(n=203) |  |  | Validation cohort<br>(n=135) |  |  | Total<br>(n=338) |
| --- | --- | --- | --- | --- | --- | --- | --- |
|  | Non-COVID<br>(n=116) | COVID<br>(n=87) | p-value | Non-COVID<br>(n=86) | COVID<br>(n=49) | p-value |  |
| Median Age (IQR), years | 68.5 (55.5-77.5) | 62 (50-75) | 0.052 | 65 (53-76) | 61 (55-75) | 0.98 | 65 (54-76) |
| sex, male | 65 (56.0%) | 41 (47.1%) | 0.21 | 49 (57%) | 23 (47%) | 0.26 | 178 (52.7%) |
| Nursing facility residence | 15 (12.9%) | 19 (21.8%) | 0.093 | 5 (6%) | 9 (18%) | 0.021 | 48 (14.2%) |
| Race |  |  |  |  |  |  |  |
| African American | 97 (83.6%) | 70 (80.5%) | 0.022 | 71 (83%) | 42 (86%) | 0.071 | 280 (82.8%) |
| Hispanic | 3 (2.6%) | 11 (12.6%) |  | 1 (1%) | 4 (8%) |  | 34 (10.1%) |
| Caucasian | 14 (12.1%) | 5 (5.7%) |  | 12 (14%) | 3 (6%) |  | 19 (5.6%) |
| Other | 2 (1.7%) | 1 (1.1%) |  | 2 (2%) | 0 (0%) |  | 5 (1.5%) |
| Smoking | 39 (33.6%) | 16 (18.4%) | 0.016 | 34 (40%) | 14 (29%) | 0.20 | 103 (30.5%) |
| Obesity | 36 (31.0%) | 49 (56.3%) | <0.001 | 33 (38%) | 27 (55%) | 0.060 | 145 (42.9%) |
| Diabetes | 51 (44.0%) | 31 (35.6%) | 0.23 | 27 (31%) | 21 (43%) | 0.18 | 130 (38.5%) |
| Hypertension | 82 (70.7%) | 53 (60.9%) | 0.14 | 51 (59%) | 32 (65%) | 0.49 | 218 (64.5%) |
| COPD | 19 (16.4%) | 9 (10.3%) | 0.22 | 11 (13%) | 4 (8%) | 0.41 | 43 (12.7%) |
| Asthma | 5 (4.3%) | 8 (9.2%) | 0.16 | 2 (2%) | 6 (12%) | 0.019 | 21 (6.2%) |
| CAD | 10 (8.6%) | 7 (8.0%) | 0.88 | 9 (10%) | 7 (14%) | 0.51 | 33 (9.8%) |
| CHF | 18 (15.5%) | 11 (12.6%) | 0.56 | 18 (21%) | 7 (14%) | 0.34 | 54 (16.0%) |
| CKD | 23 (19.8%) | 15 (17.2%) | 0.64 | 21 (24%) | 12 (24%) | 0.99 | 71 (21.0%) |
| ESRD | 13 (11.2%) | 10 (11.5%) | 0.95 | 8 (9%) | 5 (10%) | 0.86 | 36 (10.7%) |
| Atrial fibrillation | 11 (9.5%) | 4 (4.6%) | 0.19 | 4 (5%) | 7 (14%) | 0.049 | 26 (7.7%) |
| History of stroke | 14 (12.1%) | 6 (6.9%) | 0.22 | 8 (9%) | 7 (14%) | 0.38 | 35 (10.4%) |

Data are presented as median (IQR) for a continuous variable, and n (%) for categorical variables.

The dataset was split into development and validation datasets. 203 patients were assigned to the development cohort and among them 87 (42.9%) patients were tested positive for COVID-19. 135 patients were assigned to the validation cohort and among them 49 (36.3%) were tested positive. Baseline demographics and clinical characteristics of each cohort are summarized in Table 1 and Table 2.

**Table 2.** Clinical presentation and initial work-up result in the emergency room

|  | Development cohort<br>(n=203) |  |  | Validation cohort<br>(n=135) |  |  | Total<br>(n=338) |
| --- | --- | --- | --- | --- | --- | --- | --- |
|  | Non-COVID<br>(n=116) | COVID<br>(n=117) | p-value | Non-COVID<br>(n=86) | COVID<br>(n=49) | p-value |  |
| Sick contact | 4 (3.4%) | 25 (28.7%) | <0.001 | 2 (2%) | 16 (33%) | <0.001 | 47 (13.9%) |
| Triage Vital Signs |  |  |  |  |  |  |  |
| Fever | 12 (10.3%) | 25 (28.7%) | <0.001 | 9 (10%) | 11 (22%) | 0.059 | 109 (32.3%) |
| Tachycardia | 53 (45.7%) | 47 (54.0%) | 0.24 | 35 (41%) | 25 (51%) | 0.25 | 160 (47.3%) |
| Tachypnea | 42 (36.2%) | 38 (43.7%) | 0.28 | 31 (36%) | 22 (45%) | 0.31 | 133 (39.4%) |
| Hypotension | 16 (13.8%) | 17 (19.5%) | 0.27 | 8 (9%) | 5 (10%) | 0.86 | 46 (13.6%) |
| Hypoxia | 33 (28.4%) | 55 (63.2%) | <0.001 | 30 (35%) | 29 (59%) | 0.006 | 147 (43.5%) |
| Presenting symptoms |  |  |  |  |  |  |  |
| Respiratory symptoms | 60 (51.7%) | 74 (85.1%) | <0.001 | 49 (57%) | 38 (78%) | 0.016 | 221 (65.4%) |
| Cough | 31 (26.7%) | 61 (70.1%) | <0.001 | 29 (34%) | 32 (65%) | <0.001 | 153 (45.3%) |
| Shortness of breath | 54 (46.6%) | 59 (67.8%) | 0.003 | 42 (49%) | 34 (69%) | 0.021 | 189 (55.9%) |
| Constitutional symptoms | 28 (24.1%) | 55 (63.2%) | <0.001 | 27 (31%) | 24 (49%) | 0.043 | 134 (39.6%) |
| Fever | 23 (19.8%) | 47 (54.0%) | <0.001 | 15 (17%) | 24 (49%) | <0.001 | 109 (32.3%) |
| Chills | 10 (8.6%) | 21 (24.1%) | 0.002 | 10 (12%) | 9 (18%) | 0.28 | 50 (14.8%) |
| Myalgia | 7 (6.0%) | 13 (14.9%) | 0.035 | 11 (13%) | 4 (8%) | 0.41 | 35 (10.4%) |
| Chest pain | 20 (17.2%) | 15 (17.2%) | 1.00 | 14 (16%) | 9 (18%) | 0.76 | 58 (17.2%) |
| Gastrointestinal symptom | 17 (14.7%) | 33 (37.9%) | <0.001 | 20 (23%) | 21 (43%) | 0.017 | 91 (26.9%) |
| Nausea or vomiting | 11 (9.5%) | 21 (24.1%) | 0.005 | 14 (16%) | 14 (29%) | 0.090 | 60 (17.8%) |
| Diarrhea | 9 (7.8%) | 22 (25.3%) | <0.001 | 8 (9%) | 13 (27%) | 0.008 | 52 (15.4%) |
| Initial labs |  |  |  |  |  |  |  |
| Leukopenia | 5 (4.3%) | 5 (5.7%) | 0.64 | 7 (8%) | 11 (22%) | 0.019 | 28 (8.3%) |
| Leukocytosis | 32 (27.6%) | 15 (17.2%) | 0.084 | 20 (23%) | 5 (10%) | 0.060 | 72 (21.3%) |
| Thrombocytopenia | 18 (15.5%) | 19 (21.8%) | 0.25 | 10 (12%) | 11 (22%) | 0.095 | 58 (17.2%) |
| Thrombocytosis | 11 (9.5%) | 6 (6.9%) | 0.51 | 11 (13%) | 1 (2%) | 0.035 | 29 (8.6%) |
| Creatinine | 1.19<br>(0.88-1.95) | 1.12<br>(0.81-1.82) | 0.73 | 1.18<br>(0.82-2.52) | 1.33<br>(1.01-1.87) | 0.82 | 1.2<br>(0.86-1.95) |
| Initial Chest X ray |  |  |  |  |  |  |  |
| Clear lung field | 43 (40.6%) | 15 (17.9%) | <0.001 | 30 (38%) | 13 (28%) | 0.27 | 101 (32.1%) |
| Possible multifocal infiltrate | 33 (31.1%) | 43 (51.2%) | 0.005 | 31 (39%) | 25 (54%) | 0.10 | 132 (41.9%) |

Eight variables were included in the final multivariable model. Sick contact and nursing facility residence were the two biggest risk factors for COVID-19, followed by respiratory symptom (cough or shortness of breath), gastrointestinal symptom (nausea, vomiting or diarrhea), hypoxia at triage, obesity and constitutional symptom (fever, chills or myalgia). Leukocytosis was negatively associated with COVID-19 (Table 3). Based on the coefficients from the final model, a risk score was created. Sick contact and nursing facility residence were assigned two points each, respiratory symptom, gastrointestinal symptom, hypoxia at triage, obesity and constitutional symptom were assigned one point each, and leukocytosis was assigned minus one point (Table 4). A risk score of ≥ 3 achieved sensitivity of 79.3% and specificity of 80.2% in the development cohort, and sensitivity of 75.5% and specificity of 72.1% in the validation cohort. Positive and negative predictive value was 75.0% and 83.8% in the development cohort and 60.7% and 83.8% in the validation cohort, respectively. A risk scoring system for COVID-19 diagnosis achieved AUROC of 0.87 (95% CI 0.83-0.92) in the development dataset and 0.83 (95% CI 0.76-0.90) in the validation dataset (Figure 2).

**Table 3.** Results of univariable analysis and multivariable analysis

|  | Univariable analysis |  |  | Multivariable analysis |  |  |
| --- | --- | --- | --- | --- | --- | --- |
|  | OR | 95% CI | p-value | Adjusted OR | 95% CI | p-value |
| Demographics |  |  |  |  |  |  |
| Age (year) | 0.99 | 0.97-1.00 | 0.102 |  |  |  |
| Male sex | 0.70 | 0.40-1.22 | 0.209 |  |  |  |
| Nursing facility residence | 1.88 | 0.89-3.96 | 0.096 | 9.63 | 3.02-30.67 | <0.001 |
| Diabetes | 0.71 | 0.40-1.25 | 0.232 |  |  |  |
| Hypertension | 0.65 | 0.36-1.16 | 0.145 |  |  |  |
| Chronic kidney disease | 0.84 | 0.41-1.73 | 0.640 |  |  |  |
| Obesity | 2.87 | 1.61-5.11 | <0.001 | 2.93 | 1.32-6.51 | 0.008 |
| Smoking | 0.44 | 0.23-0.87 | 0.017 |  |  |  |
| Triage vital signs |  |  |  |  |  |  |
| Fever | 3.49 | 1.64-7.45 | 0.001 |  |  |  |
| Tachycardia | 1.40 | 0.80-2.44 | 0.240 |  |  |  |
| Hypotension | 1.52 | 0.72-3.21 | 0.274 |  |  |  |
| Tachypnea | 1.37 | 0.77-2.41 | 0.282 |  |  |  |
| Hypoxia | 4.32 | 2.39-7.83 | <0.001 | 3.52 | 1.58-7.83 | 0.002 |
| Sick contact | 11.29 | 3.76-33.92 | <0.001 | 10.47 | 2.67-41.04 | 0.001 |
| Presenting symptoms |  |  |  |  |  |  |
| Constitutional symptom | 5.4 | 2.94-9.93 | <0.001 | 2.31 | 1.05-5.10 | 0.038 |
| Respiratory symptom | 5.31 | 2.66-10.62 | <0.001 | 4.36 | 1.73-10.98 | 0.002 |
| Gastrointestinal symptom | 3.56 | 1.82-6.97 | <0.001 | 4.11 | 1.59-10.64 | 0.004 |
| Initial labs |  |  |  |  |  |  |
| Leukopenia |  |  |  |  |  |  |
| Leukocytosis | 0.55 | 0.27-1.09 | 0.086 | 0.33 | 0.12-0.90 | 0.03 |
| Thrombocytopenia | 1.52 | 0.74-3.11 | 0.250 |  |  |  |
| Thrombocytosis | 0.71 | 0.25-1.99 | 0.512 |  |  |  |
| Initial chest X ray |  |  |  |  |  |  |
| Clear lung field | 0.32 | 0.16-0.63 | 0.001 |  |  |  |
| Possible multifocal infiltrate | 2.32 | 1.28-4.20 | 0.005 |  |  |  |

**Table 4.** COVID-19 Risk Score at admission

|  | Points |
| --- | --- |
| Sick contact | +2 |
| Nursing Facility Residence | +2 |
| Obesity | +1 |
| Constitutional symptom (fever, chills, or myalgia) | +1 |
| Respiratory symptom (cough or shortness of breath) | +1 |
| Gastrointestinal symptom (nausea, vomiting, or diarrhea) | +1 |
| Hypoxia on triage | +1 |
| Leukocytosis | -1 |

**Figure 1.**
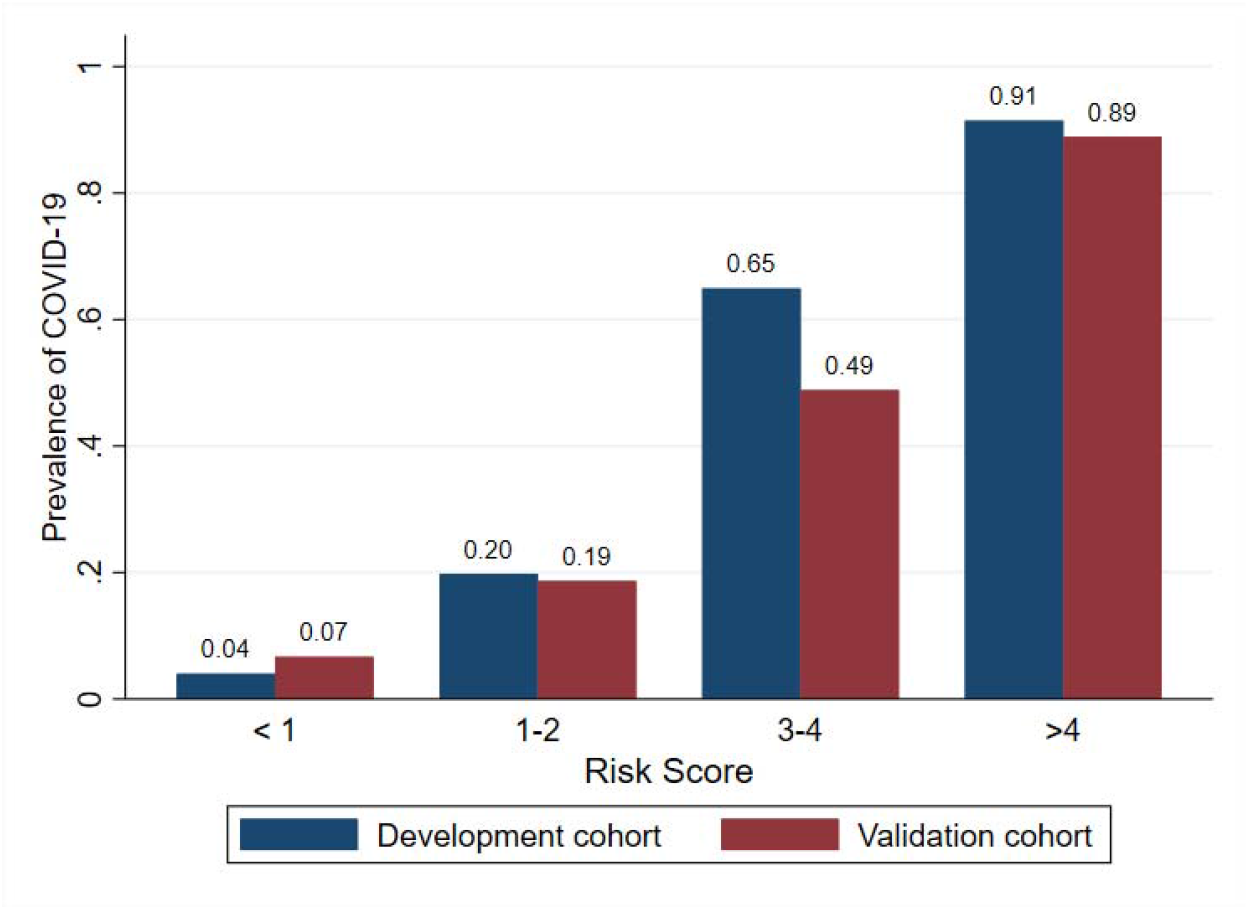
Risk score-specific prevalence rates of COVID-19

**Figure 2.**
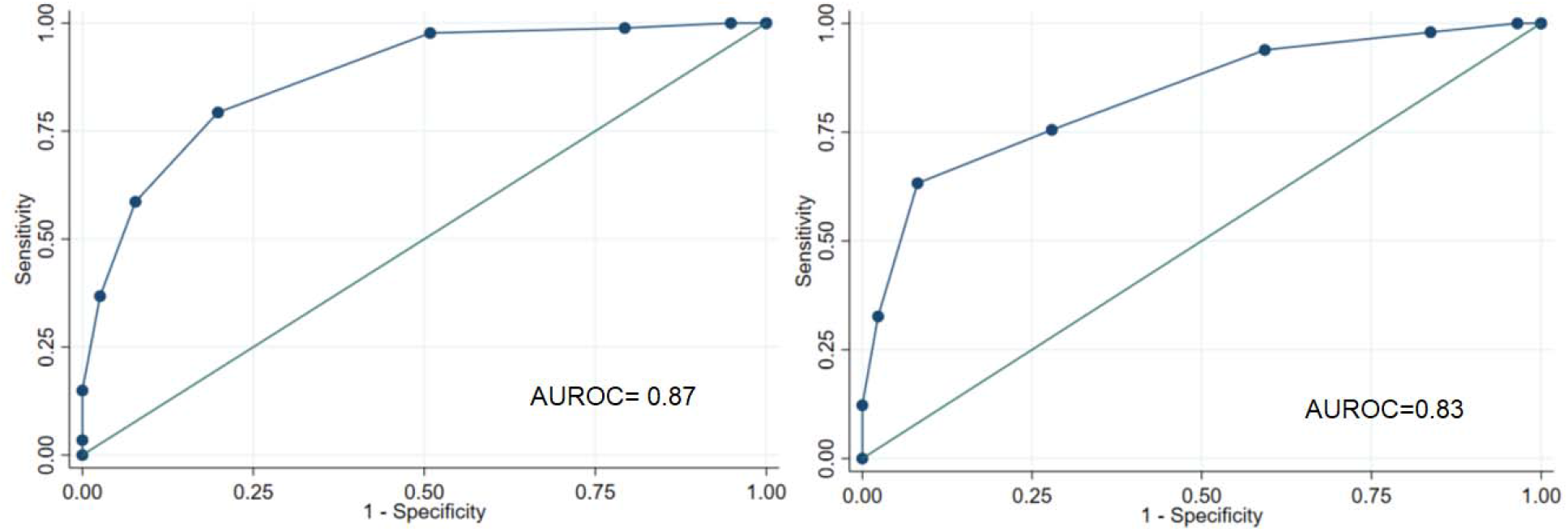
Receiver operating characteristic curve of risk score for covid-19 diagnosis among patients admitted to the hospital from the emergency room in a development cohort (left) and a validation cohort (right)

## Discussion

Early and proper identification and isolation of suspected patients with COVID-19 is essential to allow timely treatment, conserve resources, protect patients and healthcare staff and to avoid spread of COVID-19 in healthcare facilities [4]. In a resource-limited setting, it is critically important to risk-stratify patients who need testing. In this study, we present a novel bedside score developed to aid the diagnosis of COVID-19. Prior studies suggested a few logistic regression models predicting COVID-19 diagnosis [3, 5, 6]. However, logistic models are not practical to implement in clinical practice due to complex mathematical calculations needed to perform the prediction. We simplified our prediction model by creating a scoring system that is simple and practical for use, and internally validated its utility. We only included variables that are readily available at the initial hospital encounter to enable practical implementation.

In our model, sick contact and nursing facility residence were found to be two major risk factors for COVID-19 among newly admitted patients. Male sex, and chronic medical conditions such as diabetes and hypertension were known to be associated with worse outcome from COVID-19 [7-10], However, there were no significant differences in the proportion of males, hypertensives, and diabetics between COVID-19 positive admissions and negative admissions in our cohort. Obesity is another risk factor for worse clinical outcome [9, 10]. In this study, newly admitted patients with obesity were more likely to have COVID-19. In addition to demographic risk factors and symptom score, we also identified that patients with leukocytosis were less likely to have COVID-19. While COVID-19 is known to be associated with leukopenia and thrombocytopenia, we did not find significant difference in proportion of leukopenia and thrombocytopenia in cases and controls [11, 12].

In summation, the indicators used in his risk score stratification are readily available at time of admission into hospitals throughout the nation. The ability to quickly and appropriately risk stratify and identify suspected patients requiring quarantine and testing may allow physicians to make appropriate decisions in terms of early diagnosis and management.

Nonetheless, our study has limitations. Our study is limited by a small cohort size. In this study, we did not find a significant association between chest X ray findings and COVID-19 status after adjusting for the effects from confounders. Chest X ray results are likely to have clinical utility in risk-stratification of COVID-19 patients, but our study was not sufficiently powered to detect this difference. Inflammatory markers such as d-dimer, C-reactive protein, and ferritin were reported to be often elevated in COVID-19 but these lab values were not available for many study patients and therefore not included in the model [13, 14]. As our scoring system was developed in a cohort of patients admitted to the regular medical floor from the emergency room, our study result cannot be generalized to other setting, such as outpatient practices, urgent cares or intensive care units. SARS-CoV-2 is also known to cause asymptomatic infection, and our score system is designed to risk stratify newly admitted patients with symptoms concerning for COVID-19 infection, therefore cannot be used to identify asymptomatic patients. Given the above limitations and a single center study design, the risk score should be further validated in larger and/or multicenter studies.

## Conclusion

In summary, we developed a simple, easy-to-implement bedside scoring system for COVID-19 risk stratification among patients who are being admitted to the hospital. The risk score system achieved AUROC of 0.83 in validation, and can be used as a supplemental tool to assist clinical decision in the triage, quarantine, and testing of patients admitted to the hospital with suspicion of COVID-19 infection.

## Data Availability

The data for the study is available upon reasonable request with the permission of MedStar Health Research Institute.

## Acknowledgements

We thank all staff of MedStar Southern Maryland Hospital Center for their dedication and commitment to caring for the patients during the COVID-19 pandemic.

